# Reimbursement of pharmacotherapy for smoking cessation in the context of brief general practitioner advice: A cluster-randomised pilot study in the HAFO.NRW primary care research network in Germany

**DOI:** 10.1101/2025.10.31.25339228

**Authors:** Stephanie Klosterhalfen, Gabriele Franken, Zeynep Acar, Kathrin Schlößler, Ralf Jendyk, Johanna Schweizer, Larisa Pilic, Arezoo Bozorgmehr, Martina Heßbrügge, Oliver Kuß, Katharina Piedboeuf-Potyka, Susanne Kersten, Friederike Frank, Susanne Löscher, Daniel Kotz, HAFO.NRW

## Abstract

**Objective:** This study aimed to harness the potential of primary care for effective smoking cessation by piloting the effectiveness of providing free pharmacotherapy within the framework of brief general practitioner counseling.

**Methods:** In a cluster-randomised controlled pilot trial (cRCT) conducted in 2023–2024, 26 general practices from the research network HAFO.NRW were randomised. All patients received brief smoking cessation counseling by their general practitioner. Only in the intervention group was pharmacotherapy offered free of charge. The primary endpoint was self-reported tobacco abstinence after 12 weeks, biochemically validated by a carbon monoxide (CO) measurement performed in the general practice.

**Results:** A total of 129 patients from 13 intervention practices and 100 patients from 11 control practices were included. CO measurements were available for 58.1% of participants reporting abstinence. The odds ratio for CO-validated abstinence in the intervention versus control group was 1.07 (95% CI=0.25–4.58), and for self-reported abstinence 1.46 (95% CI=0.50–4.26). More patients in the intervention group used nicotine replacement therapy to support their quit attempt (81.4% vs. 56.1% in the control group), while more patients in the control group used e-cigarettes (27.3% vs. 17.6% in the intervention group).

**Conclusion:** Offering free pharmacotherapy in primary care appears feasible and may encourage the use of evidence-based cessation aids. These findings warrant further investigation in a confirmatory trial.

## INTRODUCTION

Smoking is the leading preventable cause of disease and death and a major risk factor for cancer, cardiovascular disease, and respiratory illness (1,2). In Germany, around 125,000 people die each year as a result of smoking (3), with 28% of these deaths occurring among people of working age (3). The economic costs are estimated at around 97 billion euros per year, including 30 billion euros in direct healthcare costs (4).

Despite these figures, smoking remains widespread in Germany: in 2024, 29% of the population smoked – 32% of men and 26% of women (5). Smoking is particularly prevalent among socioeconomically disadvantaged groups, where the chances of successfully quitting are also lower (6,7).

Nicotine, contained in cigarettes, is highly addictive (8). Without support, fewer than 5% of smokers manage to quit (9). The German S3 guideline on the treatment of tobacco dependence therefore recommends a combination of brief medical counselling and pharmacotherapy, such as nicotine replacement therapy (NRT) or prescription medication (10). One particularly efficient form of brief medical counselling is the ABC method *(Ask, Brief Advice, Cessation support)* (11).

International studies show that when the costs of cessation aids are covered, both their use and the success rates of quitting increase (12–14). Although around 68% of smokers in Germany regularly use general medical services, only 1.6% report having received medical advice on quitting combined with a recommendation for pharmacotherapy (15).

The Health Care Further Development Act (GVWG, 2021) established the basis for cost coverage by statutory health insurance. The IQWiG (Institute for Quality and Efficiency in Health Care) has already issued a positive assessment of pharmacological cessation treatments (16). Severely nicotine-dependent insured individuals are now entitled, once, to pharmacy-only cessation medication as part of evidence-based programmes (17). The corresponding decision by the Federal Joint Committee (G-BA) came into force on 20 August 2025. Previously, the lack of cost coverage was considered a major barrier – GPs were less likely to recommend such measures, and financial obstacles could discourage individuals from attempting to quit.

This cluster-randomised controlled pilot study, conducted within the HAFO.NRW primary care research network; www.hafo.nrw), examined whether providing guideline-based pharmacotherapy for smoking cessation free of charge, in combination with brief GP counselling, increases tobacco abstinence after 12 weeks compared to standard care.

In addition, the study explored whether the intervention influenced smoking cessation attempts (including the number of attempts) and to what extent supportive measures were used. The results are intended to provide initial insights into whether offering cessation medication free of charge could be an effective strategy for reducing tobacco use in Germany.

## METHODS

The study was conducted as part of HAFO.NRW, a collaborative project funded by the Federal Ministry of Education and Research (BMBF) to establish a sustainable research infrastructure in primary care. The network includes the general practice departments of the eight medical faculties in North Rhine-Westphalia (Aachen, Bochum, Bonn, Düsseldorf, Essen, Cologne, Münster, and Witten/Herdecke) and integrates general practices as “research practices” into applied, epidemiological, and clinical studies. In these practices, general practitioners (GPs) and medical assistants (MAs) work in tandem with universities to address practice-relevant research questions and to strengthen general practice as an academic discipline.

### Study design

This study was designed as a cluster-randomised controlled pilot trial (cRCT). Its aim was to evaluate the feasibility and potential impact of providing guideline-based pharmacotherapy for smoking cessation free of charge within a primary care setting. To prevent contamination within practices and to test the practical applicability of the intervention, randomisation was carried out at the practice (cluster) level.

A detailed, pre-registered study protocol is available at https://osf.io/txwer. Blinding of participants was not possible; however, the analysis plan was finalised and publicly registered prior to inclusion of the first participant. The data analysts were blinded to group allocation.

The Coordinating Centre for Clinical Trials (KKS) Düsseldorf was responsible for quality assurance and monitoring in accordance with Good Clinical Practice (GCP). As part of this, one monitoring visit was conducted in each participating practice.

All participating GPs and MAs received a handout detailing the study procedures. In addition, GPs took part in an established training session on the ABC method. This online blended-learning format included a two-hour interactive live webinar with peer coaching by an experienced GP and exercises with simulated patients. Preparatory materials included a screencast on tobacco dependence and evidence-based cessation methods, as well as a video demonstrating best practices in the ABC method. Participants also received one-page handouts summarising the core principles of the ABC approach and available treatment options.

### Inclusion and exclusion criteria

Eligible practices were members of the HAFO.NRW research network. Practices directly affiliated with any HAFO staff member (e.g., through ownership or employment) were excluded. Eligible patients were 18 years or older and smoked at least 10 cigarettes per day. Exclusion criteria included pregnancy or breastfeeding, moderate to severe cognitive impairment, and insufficient German language skills.

### Intervention group

Patients received brief GP counselling on smoking cessation and were offered the option of cost-free pharmacotherapy, decided jointly with their GP. The medication could be collected from a pharmacy, with costs covered by HAFO.NRW. Billing was handled independently of the patient, mirroring a scenario close to standard health insurance procedures.

The treatment duration was up to 12 weeks, with follow-up prescriptions possible. GPs could invite patients for a follow-up consultation within a week, and additional sessions could be scheduled as needed to provide behavioural support and encourage adherence.

All patients completed questionnaires at baseline and 12-week follow-up (either online or on paper). Baseline data included smoking status, consumption patterns, dependence level, and motivation to quit; follow-up data focused on quit attempts and cessation methods used.

### Control group

The procedure was identical to that of the intervention group, except those patients had to pay for cessation pharmacotherapy themselves.

### Primary and secondary endpoints

Following international recommendations for evaluating smoking cessation interventions (18) the primary endpoint was self-reported abstinence at 12 weeks, biochemically validated by carbon monoxide (CO) measurement in exhaled air (cut-off < 8 ppm to confirm abstinence).

Each practice was equipped with a CO measuring device (Smokerlyzer) and trained in its use. Patients without CO validation were classified as smokers, in line with international standards for smoking cessation trials(19).

Secondary endpoints included quit attempts and the use of pharmacotherapy or other support measures, assessed via the follow-up questionnaire after approximately 12 weeks.

### Sample size

The target sample size was 288 patients, enrolled across 12 intervention and 12 control practices (12 patients per practice). Considering the cluster design (intraclass correlation coefficient [ICC] = 0.05 (20), design effect = 1.55), the study had a moderate power (57%) to detect an abstinence rate of 24% in the intervention group versus 12% in the control group, at a significance level of α = 5%(11,21,22).

### Randomisation

Practices were enrolled sequentially between May 2023 and February 2024, participating in one of six training cycles before randomisation. The randomisation sequence was generated using SAS PROC PLAN with variable block sizes of 2, 4, or 6 to ensure allocation concealment at the cluster level (practice). No stratification or a priori matching of practices was applied.

### Statistical analysis

Primary (CO-validated abstinence) and secondary outcomes were compared between groups using logistic regression, incorporating a random intercept to account for clustering at the practice level.

The primary analysis was conducted on an intention-to-treat basis. Three sensitivity analyses were performed to test robustness: (1) Based solely on self-reported abstinence. (2) Adjusted for potential confounders (time between baseline and follow-up, sex, age, level of nicotine dependence, and motivation to quit). (3) Based on self-reported abstinence, also adjusted for the above covariates.

## RESULTS

A total of 26 general practices were enrolled and evenly randomised into the intervention and control groups (see Figure 1). After randomisation, two control practices withdrew due to staff restructuring. The remaining 24 practices initially recruited 238 patients (134 in the intervention and 104 in the control group). Before analysis, it was determined that five patients in the intervention group and four in the control group did not meet the inclusion criterion of smoking at least ten cigarettes per day. These were excluded, leaving 229 patients (129 in the intervention group) in the final analysis.

**Figure. 1.**
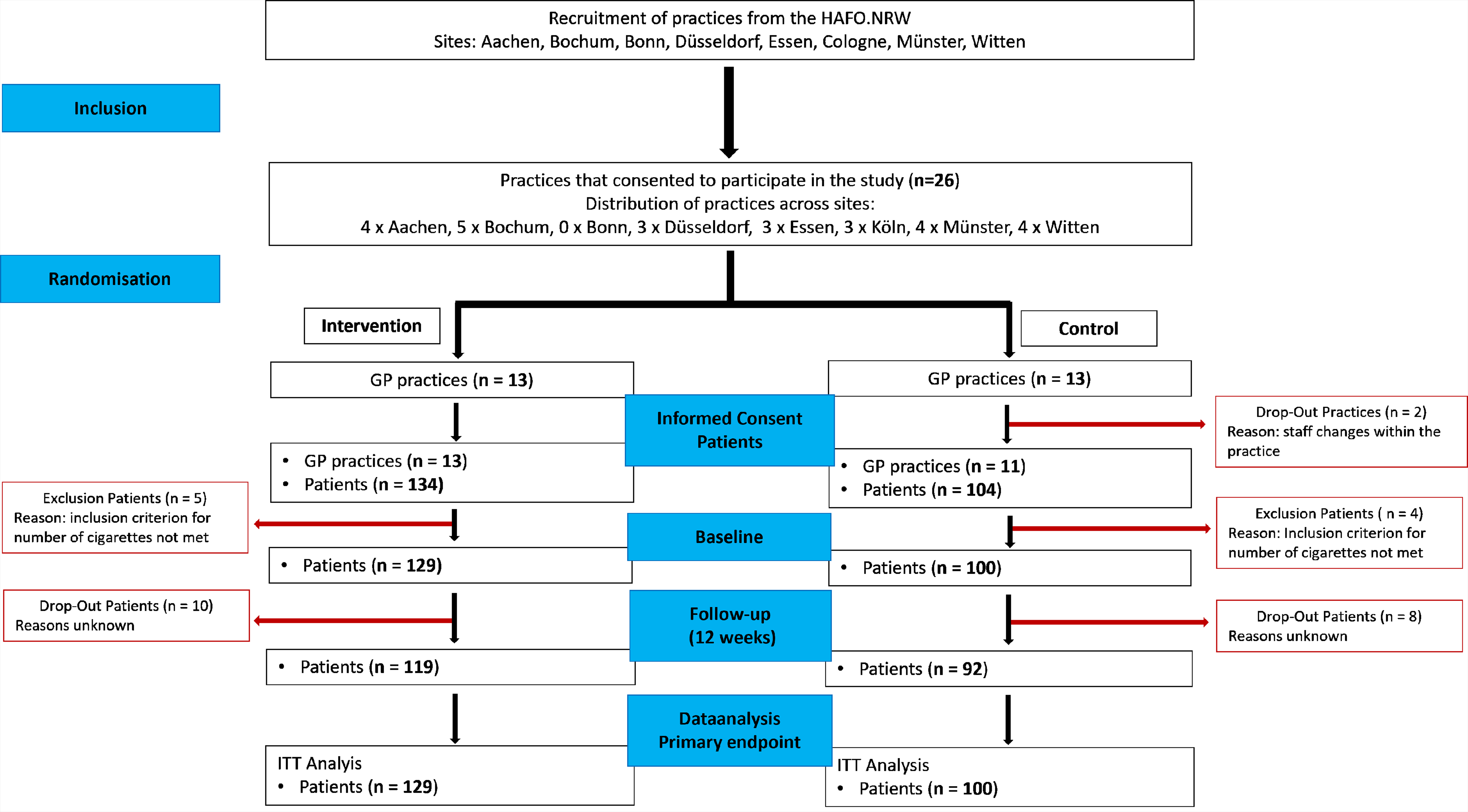

Table 1 presents the baseline characteristics of participants. The mean age was 53.3 years (SD = 14.2) in the intervention group and 50.6 years (SD = 14.2) in the control group. Patients in the intervention group smoked more cigarettes per day and showed a higher proportion with moderate to high dependence. Motivation to quit was similar across groups, though a slightly higher proportion of participants in the intervention group reported high motivation (38.8% vs 31.0%).

**Table 1:** Sociodemographic and smoking-related characteristics of participants at baseline by study group.

| Characteristics | Intervention<br>n = 129 (from 13 GP<br>practices) | Control<br>n = 100 (from 11 GP practices) |
| --- | --- | --- |
| Sex, % (n) |  |  |
| Male | 48.8 (63) | 44.0 (44) |
| Female | 50.4 (65) | 55.0 (55) |
| N/A | 0.8 (1) | 1.0 (1) |
| Age, Median (SD) | 53.3 (14.2) | 50.6 (14.2) |
| Cigarettes per day, Median(SD) | 23.7 (9.6) | 20.6 (4.9) |
| Cigarette dependence<sup>~</sup>, % (n) |  |  |
| High | 5.4 (7) | 8.0 (8) |
| Medium | 79.8 (103) | 69.0 (69) |
| Low | 14.7 (19) | 23.0 (23) |
| Motivation to stop smoking<sup>#</sup>, % (n) |  |  |
| High | 38.8 (50) | 31.0 (31) |
| Medium | 58.9 (76) | 63.0 (63) |
| Low | 2.3 (3) | 6.0 (6) |

<sup>~</sup> Measured with „Heaviness of Smoking Index“ (HSI) (27,28). Dependence is calculated based on the number of cigarettes smoked per day and the time to the first cigarette after waking up: Low = level 0-1, medium = level 2-4, high = level 5-6;
<sup>#</sup> Motivation to Stop Smoking Scale (29,30): Low = level 0-1, medium = level 2-5, high = level 6-7.

The mean interval between baseline and follow-up was around 13 weeks (94 days) in the intervention group and 14 weeks (99 days) in the control group, slightly longer than the planned 12 weeks (84 days). At follow-up, 21.7% (28/129) of patients in the intervention group reported tobacco abstinence, with CO-validated abstinence confirmed in 9.3% (12/129). In the control group, 15.0% (15/100) reported abstinence, with CO-validated abstinence in 9.0% (9/100).

CO measurement for objective validation of self-reported abstinence was not performed consistently. In the intervention group, 28 participants reported abstinence; of these, 16 had documented CO measurements, and 12 provided measurable values – all confirming abstinence (12/12, 100%). In the control group, 15 participants reported abstinence; 13 had CO readings, but 4 exceeded 8 ppm, and thus were not validated as abstinent. Validated abstinence was therefore achieved in 69.2% (9/13) of control participants. Overall, 25 of 43 self-reported abstainers (58.1%) had documented CO measurements.

Analysis showed an odds ratio (OR) for CO-validated abstinence (= primary endpoint) of 1.07 (95% CI 0.25–4.58; p = 0.93). Sensitivity analyses yielded an OR of 1.46 (95% CI 0.50–4.26; p = 0.49) for self-reported abstinence. Adjusted models produced comparable estimates (see Table 2).

**Table 2:** Results of logistic regression analyses for tobacco abstinence – unadjusted and adjusted models accounting for covariates.

| Analysis | Intervention vs. Control |  |
| --- | --- | --- |
|  | OR (95%-CI) | p |
| **CO-validated abstinence (primary endpoint)** | 1.07 (0.25-4.58) | 0.929 |
| **Sensitivity analyses** |  |  |
| 1) Self-reported abstinence | 1.46 (0.50-4.26) | 0.487 |
| 2) Adjusted~: CO-validated abstinence | 0.81 (0.18-3.71) | 0.785 |
| 3) Adjusted~: self-reported abstinence | 1.26 (0.45-3.54) | 0.666 |

~ Adjusted for the following covariates: duration in days between baseline and follow-up, sex, age, level of cigarette dependence, and motivation to quit smoking.

A quit attempt was reported by 92.4% (110/119) of patients in the intervention group and 81.5% (75/92) in the control group; the average number of attempts was slightly higher in the intervention group (see Table 3). The most frequently reported cessation method was GP counselling (85.3% vs 86.4%). Use of nicotine replacement therapy (NRT) was higher in the intervention group (81.4% vs 56.1%), while e-cigarette use was lower (17.6% vs 27.3%; see Table 3).

**Table 3:** Self-reported quit attempts and cessation methods of patients at follow-up.

|  | Patients follow-up |  |
| --- | --- | --- |
|  | Intervention<br>n = 119 | Control<br>n = 92 |
| **Quit attempts** | **% (n)** | **% (n)** |
| Patients with at least on quit attempt, % (n) | 92.4 (110) | 81.5 (75) |
| Number of quit attempts (mean, SD) | 3.2 (9.2) | 2.2 (1.8) |
| **Methods<sup>~</sup>** | **Intervention**<br>**(n = 102)** | **Control**<br>**(n = 66)** |
| GP counselling | 85.3 (87) | 86.4 (57) |
| Specialist counselling (excluding GP) | 2.0 (2) | 3.0 (2) |
| Smoking cessation courses / individual therapy | 7.8 (8) | 1.5 (1) |
| Telephone counselling (e.g., quitline) | 1.0 (1) | 0.0 (0) |
| Nicotine replacement therapy (e.g., nicotine gum) | 81.4 (83) | 56.1 (37) |
| Medication containing bupropion | 4.9 (5) | 9.1 (6) |
| Medication containing varenicline | 0.0 (0) | 0.0 (0) |
| Medication containing cytisine | 0.0 (0) | 0.0 (0) |
| E-cigarette | 17.6 (18) | 27.3 (18) |
| Heated tobacco product | 4.9 (5) | 6.1 (4) |
| Smoking cessation app | 4.9 (5) | 3.0 (2) |
| Smoking cessation website | 7.8 (8) | 4.5 (3) |
| Book on smoking cessation | 8.8 (9) | 7.6 (5) |
| Hypnotherapy | 2.9 (3) | 0.0 (0) |
| Acupuncture | 1.0 (1) | 3.0 (2) |
| Alternative practitioner | 2.9 (3) | 1.5 (1) |
| Other method not listed above | 3.9 (4) | 4.5 (3) |

<sup>~</sup>List of response options according to the follow-up questionnaire – multiple answers possible; light blue shading: the three most frequently used cessation methods

Table 4 summarises GP prescribing behaviour. Data were available from 12 intervention practices (123 patients) and 7 control practices (65 patients). Nicotine patches and gums were the most frequently prescribed treatments in both groups, but overall, NRT was recommended more often in the intervention group, particularly nicotine patches (81.3% vs 46.2%).

**Table 4:** Prescriptions of pharmacotherapy for tobacco abstinence as reported by general practices.

| Methods<sup>~</sup> | Intervention<br>n= 123 (from 12/13 GP<br>practices)<sup>#</sup> | Control<br>n = 65 (from 7/11 GP<br>practices)<sup>#</sup> |
| --- | --- | --- |
|  | % (n) | % (n) |
| Nicotine replacement therapy (NRT) |  |  |
| Nicotine patch | 81.3 (100) | 46.2 (30) |
| Nicotine gum | 35.0 (43) | 16.9 (11) |
| Nicotine spray | 25.2 (31) | 3.1 (2) |
| Nicotine lozenge | 13.0 (16) | 4.6 (3) |
| Medication containing bupropion | 8.9 (11) | 6.2 (4) |

<sup>~</sup>Multiple answers possible; <sup>#</sup> Data on prescribed pharmacotherapy were not available from all practices.

A post hoc cost analysis based on pharmacy billing data (see Supplementary Table 1) showed the highest total costs for nicotine patches (∼€17,735), followed by gums (∼€3,809) and lozenges (∼€1,889). The mean cost per patient was approximately €197.

## DISCUSSION

This pilot study indicates that covering the cost of pharmacotherapy in primary care can encourage the use of cessation medications. Brief GP counselling combined with pharmacological support proved feasible and resulted in greater use of pharmacotherapy, particularly nicotine replacement. Although the odds ratio for CO-validated abstinence was only slightly higher and not statistically significant, patients in the intervention group reported more quit attempts and greater use of cessation aids – suggesting that cost-free access may promote both utilisation and quit attempts.

The average treatment cost of approximately €197 per patient is low compared with the potential long-term healthcare costs of smoking-related diseases, highlighting the economic viability of such an approach.

International studies have shown that providing free pharmacotherapy in primary care settings increases treatment use – especially among socioeconomically disadvantaged groups – and improves abstinence rates (23–25). Similarly, in this study, more quit attempts and slightly higher self-reported abstinence were observed in the intervention group, although without statistical significance.

The sample size calculation assumed abstinence rates of 24% in the intervention group and 12% in the control group; observed self-reported abstinence was 22% vs 15%. However, CO validation proved challenging: in the ITT analysis, validated abstinence rates were 9.3% vs 9%, partly due to missing CO data in the intervention group, which were classified as non-abstinent. Baseline group differences must also be considered – particularly the higher nicotine dependence in the intervention group, which likely reduced their baseline probability of successful cessation. The low rate of CO validation and non-adherence to inclusion criteria highlight implementation issues to be addressed in a future confirmatory RCT. Nevertheless, the higher number of quit attempts and greater use of evidence-based NRT suggest a potential intervention effect.

Another key finding is the overall low uptake of pharmacotherapy – a result consistent with previous research, such as a Dutch RCT (26). Despite full cost coverage, only a subset of patients opted for pharmacological support. Possible reasons include low perceived need, concerns about side effects, or a preference for non-pharmacological approaches (by either GPs or patients). Earlier studies indicate that even in countries with full reimbursement, additional measures such as intensive counselling or targeted education campaigns are needed to increase uptake (14).

An additional finding was that offering evidence-based pharmacotherapy (NRT) in the intervention group was associated with lower use of e-cigarettes, which are not recommended in clinical guidelines for smoking cessation (10).

### Strengths and Limitations

Strengths of this study include its cluster-randomised design, pragmatic implementation in real-world primary care with pharmacy involvement, and CO-validated measurement of the primary endpoint. The structured ABC training for GPs and standardised intervention protocol strengthen internal validity.

Limitations include missing or incomplete CO measurements despite self-reported abstinence, indicating implementation challenges that limited objective validation. Future studies should use standardised procedures and quality control to improve data completeness. Potential selection bias in patient recruitment – for instance, inclusion of more highly dependent smokers in intervention practices – was accounted for in sensitivity analyses. Self-selection of particularly motivated practices may also limit generalisability.

Furthermore, baseline imbalances existed between groups, particularly in daily cigarette consumption, dependence level, and motivation to quit, which may have affected comparability and must be considered when interpreting results.

A larger, multicentre study with extended follow-up would be useful to assess the long-term effects of cost coverage on smoking prevalence.

## Data Availability

All data produced in the present study are available upon reasonable request to the authors

## REGISTRATION

The study was approved by the Ethics Committee of the Medical Faculty of Heinrich Heine University Düsseldorf (reference: 2022-2107) and registered in the German Clinical Trials Register (DRKS00031777).

## ACKNOWLEDGEMENTS

Our sincere thanks go to all participating general practitioners, medical assistants, and patients, as well as to the Coordinating Centre for Clinical Trials Düsseldorf (KKSD) for their support.

---

**Supplementary Table 1:** Pharmacy-submitted invoices for pharmacotherapy for tobacco abstinence.

| Methods | Frequency | Costs |
| --- | --- | --- |
|  | % (n) | € |
| **Nicotine replacement therapy (NRT)** |  |  |
| Nicotine patch | 69.8 (400) | 17,735.11 |
| Nicotine gum | 16.2 (94) | 3,808.99 |
| Nicotine spray | 4.0 (23) | 1,153.01 |
| Nicotine lozenge | 9.0 (52) | 1,889.22 |
| **Medication** containing bupropion | 2.0 (11) | 831.87 |
| **Cost per patient (average)** |  | **197.04** |

Data on submitted invoices for pharmacotherapy were available, due to the study design, from 13 pharmacies for the intervention group comprising 129 patients.

